# Clinical features and burden of post-acute sequelae of SARS-CoV-2 infection in children and adolescents: an exploratory EHR-based cohort study from the RECOVER program

**DOI:** 10.1101/2022.05.24.22275544

**Authors:** Suchitra Rao, Grace M. Lee, Hanieh Razzaghi, Vitaly Lorman, Asuncion Mejias, Nathan M. Pajor, Deepika Thacker, Ryan Webb, Kimberley Dickinson, L. Charles Bailey, Ravi Jhaveri, Dimitri A. Christakis, Tellen D. Bennett, Yong Chen, Christopher B. Forrest

## Abstract

**Importance:** The post-acute sequelae of SARS-CoV-2 (PASC) has emerged as a long-term complication in adults, but current understanding of the clinical presentation of PASC in children is limited.

**Objective:** To identify diagnosed symptoms, diagnosed health conditions and medications associated with PASC in children.

**Design, Setting and Participants:** Retrospective cohort study using electronic health records from 9 US children’s hospitals for individuals <21 years-old who underwent reverse transcriptase polymerase chain reaction (RT-PCR) testing for SARS-CoV-2 between March 1, 2020 – October 31, 2021 and had at least 1 encounter in the 3 years before testing.

**Exposure:** SARS-CoV-2 PCR positivity.

**Main Outcomes and Measures:** We identified syndromic (symptoms), systemic (conditions), and medication PASC features in the 28-179 days following the initial test date. Adjusted hazard ratios (aHRs) were obtained for 151 clinically predicted PASC features by contrasting PCR-positive with PCR-negative groups using proportional hazards models, adjusting for site, age, sex, testing location, race/ethnicity, and time-period of cohort entrance. We estimated the incidence proportion for any syndromic, systemic or medication PASC feature in the two groups to obtain a burden of PASC estimate.

**Results:** Among 659,286 children in the study sample, 59,893 (9.1%) tested positive by PCR for SARS-CoV-2. Most were tested in outpatient testing facility (50.3%) or office (24.6%) settings. The most common syndromic, systemic, and medication features were loss of taste or smell (aHR 1.96 [95% CI 1.16-3.32), myocarditis (aHR 3.10 [95% CI 1.94-4.96]), and cough and cold preparations (aHR 1.52 [95% CI 1.18-1.96]). The incidence of at least one systemic/syndromic/medication feature of PASC was 41.9% among PCR-positive children versus 38.2% among PCR-negative children, with an incidence proportion difference of 3.7% (95% CI 3.2-4.2%). A higher strength of association for PASC was identified in those cared for in the ICU during the acute illness phase, children less than 5 years-old, and individuals with complex chronic conditions.

**Conclusions and Relevance:** In this large-scale, exploratory study, the burden of pediatric PASC that presented to health systems was low. Myocarditis was the most commonly diagnosed PASC-associated condition. Acute illness severity, young age, and comorbid complex chronic disease increased the risk of PASC.

**Key Points:** *Question:* What are the incidence and clinical features of post-acute sequelae of SARS-CoV-2 infection (PASC) in children?

*Findings:* In this retrospective cohort study of 659,286 children tested for SARS-CoV-2 by polymerase chain reaction (PCR), the symptom, condition and medication with the strongest associations with SARS-CoV-2 infection were loss of taste/smell, myocarditis, and cough and cold preparations. The incidence proportion of non-MIS-C related PASC in the PCR-positive group exceeded the PCR-negative group by 3.7% (95% CI 3.2-4.2), with increased rates associated with acute illness severity, young age, and medical complexity.

*Meaning:* PASC in children appears to be uncommon, with features that differ from adults.

## Introduction

Scientific and clinical evidence is evolving on the sequelae following coronavirus disease 2019 (COVID-19) caused by Severe Acute Respiratory Syndrome Coronavirus 2 (SARS-CoV-2). Post-acute sequelae of SARS-CoV-2 (PASC) infection has a broad spectrum of clinical manifestations that can affect multiple organ systems. The National Institutes of Health definition of PASC refers to ongoing, relapsing or new symptoms or other health effects occurring after the acute phase of SARS-CoV-2 infection that is present four or more weeks after the acute infection.^1^ As defined by the Centers for Disease Control and Prevention, post-COVID-19 conditions include a wide range of new, recurring, or persistent health problems people experience four or more weeks after being infected with SARS-CoV-2.^2^ Alternatively, the World Health Organization defines post-COVID-19 conditions as those occurring ≥3 months from the onset of SARS-CoV-2 infection that cannot be explained by an alternative diagnosis, and last for at least 2 months, acknowledging that a separate definition may be applicable for children.^3^

In adults, PASC is characterized by two types of manifestations: persistent, intermittent or relapsing non-specific symptoms such as fatigue, headache and shortness of breath (a syndromic variant) and conditions which may be a direct result of the acute illness presentation (such as pulmonary fibrosis) or that may arise *de novo* after infection, such as autoimmune conditions (a systemic variant).^4^ While more data are accruing in adults,^5-12^ our understanding of PASC is still limited in children, with the exception of MIS-C.^13^ There is an urgent need to elucidate the incidence, clinical features and duration of PASC in children, with standardized definitions and data collection methods.^14^

There is wide variability in prevalence estimates of PASC in children ranging from 2-66%, due to the marked heterogeneity in case ascertainment and methodology of existing studies, as well as lack of an adequate comparison group.^15-20^ It remains unclear to what extent children experience SARS-CoV-2 sequelae, and how PASC features may differ from those in adults. In this study our objectives were to 1) identify syndromic (symptoms) and systemic (conditions) features, and medications used to treat PASC in the 1-6 months following SARS-CoV-2 infection in children and adolescents, 2) estimate the incidence of any PASC feature attributable to SARS-CoV-2 infection, and 3) to identify risk factors for PASC.

## Methods

### Data Source

This retrospective cohort study is part of the NIH Researching COVID to Enhance Recovery (RECOVER) Initiative, which seeks to understand, treat, and prevent the post-acute sequelae of SARS-CoV-2 infection (PASC). For more information on RECOVER, visit https://recovercovid.org/. We used electronic health record (EHR) data from PEDSnet institutions for the study. PEDSnet (pedsnet.org) is a multi-institutional clinical research network that aggregates EHR data from several of the nation’s largest children’s healthcare organizations.^21,22^ Participating institutions included Children’s Hospital of Philadelphia, Cincinnati Children’s Hospital Medical Center, Children’s Hospital Colorado, Ann & Robert H. Lurie Children’s Hospital of Chicago, Nationwide Children’s Hospital, Nemours Children’s Health System (a Delaware and Florida health system), Seattle Children’s Hospital, and Stanford Children’s Health. PEDSnet standardizes institutional data to the Observational Medical Outcomes Partnership common data model. PEDSnet has accrued data for >8 million children with at least one visit from 2009-2021 from inpatient and outpatient clinical settings.

We retrieved EHR data from all healthcare encounters associated with patients who underwent SARS-CoV-2 polymerase chain reaction (PCR) testing provided in outpatient, inpatient and emergency department settings. Data were extracted from the PEDSnet COVID-19 Database-Version 2022-01-16 on January 25, 2022 and included EHR data with dates of services up to December 31, 2021. The Children’s Hospital of Philadelphia’s Institutional Review Board designated this study as not human subjects’ research and waived informed consent. Reporting of study design and results follows the Strengthening the Reporting of Observational Studies in Epidemiology (STROBE) reporting guideline for observational research.^23^

### Cohorts

Infected cohorts were individuals < 21 years of age at the time of the health encounter with a positive SARS-CoV-2 PCR test between March 1, 2020 and October 31, 2021. We compared this group to a cohort of children with negative SARS-CoV-2 PCR (and no prior or subsequent positive) tests during the same study period. Children with a positive serology test, or MIS-C/PASC diagnosis codes with negative PCR tests were also excluded from the comparison group. Cohort entry was defined by the date of the first positive (infected) or first negative test. We further restricted the sample to those with at least one encounter (including telephone/telehealth) within health systems from day 7 to day 1095 before cohort entry to ensure prior care at a PEDSnet institution.

### Outcomes

We developed International Classification of Diseases-10-CM (ICD-10) syndromic (symptoms), systemic (health conditions), and medication (therapeutic classes) clusters of codes, which served as outcomes. We coded 66,294 ICD-10-CM codes into 439 deduplicated clusters, beginning with the widely used Agency for Healthcare Research and Quality (AHRQ) Clinical Classification Software Refined,^24^ and extending the categories to include more refined autoimmune, mental health disorders, and COVID-19-related complication categories. ICD codes were obtained from all service settings including telephone encounters, inpatient, outpatient clinics, and emergency departments. Syndromic clusters included symptoms such as fever, cough, fatigue, shortness of breath, chest pain, palpitations, chest tightness, headache, and altered smell and taste. Systemic clusters included such conditions as multi-inflammatory syndrome, myocarditis, diabetes, and other autoimmune diseases. We clustered 2,172 RxNorm medication ingredient codes into 228 Anatomical Therapeutic Chemical (ATC) Classification Level 3 classes for prescribed medications that were listed in the record during the outcome evaluation period.^25^ Examples include antiarrhythmics, opioids, and systemic corticosteroids. The full list of the code assignments to clusters is publicly available (https://github.com/PEDSnet/PASC).

To evaluate their clinical content validity, all clusters were reviewed by experts in pediatric psychiatry, infectious diseases, cardiology, pulmonology, and primary care. We narrowed this list to *an a priori* set of 121 syndromic and systemic and 30 medication clusters that were clinically predicted to be PASC-associated based on our review of the literature, clinical experience, and potential medications used to treat PASC-associated symptoms and conditions.

The outcome assessment follow-up time period spanned 28 to 179 days after cohort entrance, or up to December 31, 2021, whichever event came first. For chronic conditions (e.g., asthma, diabetes, etc.), we excluded patients from the denominator if they had evidence of that condition in the 18 months before cohort entrance.

### Covariates

Age was assessed at cohort entry. Other covariates were sex, race/ethnicity, institution, obesity, testing location (emergency department, inpatient, outpatient clinic, or outpatient testing facility), intensive care unit care 7 days before through 13 days after cohort entrance, and medical complexity. Obesity was defined as presence of an age-sex standardized BMI z-score ≥95^th^ percentile any time before cohort entrance. We used the Pediatric Medical Complexity Algorithm (PMCA) Version 2.0^26^ to categorize children as having no chronic condition, non-complex chronic condition, or complex chronic conditions. We considered diagnoses up to three years before cohort entrance. Children were assigned to the complex chronic condition category if they had conditions that affected ≥2 body systems (e.g., Type 1 diabetes with end-organ complications) or a progressive chronic condition (e.g., muscular dystrophy).^27^

### Statistical Analyses

We used descriptive analyses to report characteristics of tested patients, divided into PCR-positive and PCR-negative patients, and contrasted the groups using standardized differences^28^ and set a cut-point of >0.20 as a meaningful difference. To explore post-acute sequelae of SARS-CoV-2 infection, we evaluated each clinically predicted symptom, condition, and medication as a separate outcome for a total of 121 syndromic/systemic clusters and 30 classes of medications. This was done using Cox proportional hazards models, adjusting for institution, age group (<1 year, 1-4 years, 5-11 years, 12-15 years and 16-< 21 years), sex, race/ethnicity, time period of cohort entrance (March-June 2020, July-October 2020, November 2020-February 2021, March-June 2021, July-Oct 2021), location of testing (ED, inpatient, outpatient office, outpatient test only, other/unknown), and medical complexity. Each regression model produced an adjusted hazard ratio (aHR) and 95% confidence interval that contrasted the risk (i.e., incidence) of the feature in the PCR-positive cohort with the PCR-negative cohort. aHRs for which the 95% CI lower limit was >1.00 were considered to be empirically supported. We displayed these aHRs in forest plots and used this subset in further analyses. This approach permitted evaluation of all encounters in the outcome evaluation period, thus minimizing the risk of bias that may result from children returning from care within the health system, versus those who did not. We implemented a false discovery rate adjustment for our p-values and 95% confidence intervals in our forest plots. This multiple-testing adjustment procedure controls for the proportion of false coverage statements expressed by the reported confidence intervals to be less than 0.05.^29^

To estimate the incidence of non-MIS-C PASC, we excluded children with MIS-C diagnoses and calculated the difference in the incidence proportions for any clinically-predicted and empirically-supported syndromic, systemic or medication-related PASC feature between children in the SARS-CoV-2 PCR-positive group and those with at least one PASC feature in the SARS-CoV-2 test negative group.

We calculated the standardized morbidity ratio, by applying the age-viral testing date-specific PASC incidence rates (ie, 25 different rates) for the PCR-negative group to the PCR-positive group to obtain expected counts and contrasted those with the number observed.

Finally, we evaluated the risk of any syndromic, systemic or medication feature, by age, comorbidity and illness severity to identify features associated with the risk of non-MIS-C PASC. Analyses were conducted using R version 4.1.2 (2021-11-01).

## Results

### Cohort identification

From March 1, 2020 to October 31, 2021, there were 1,782,537 patients < 21 years of age, of which 817,505 (45.9%) patients were tested for SARS-CoV-2 by PCR across all sites; of these, 659,286 had at least one visit in the 7-1095 days before the PCR-test date. Of this total, 59,893 tested positive (9.1%). The follow-up time during the outcome assessment period for both positive and negative groups were similar (mean 4.6 months versus 4.7 months). Most patients were tested in outpatient testing facilities (50.3%) or outpatient clinic (24.6%) settings. PCR-positive compared with PCR-negative patients were more likely to be Non-Hispanic Black/African-American (20.2% vs 15.4%), Hispanic ethnicity (18.8% vs 15.3%), and older (17.4% vs. 12.1% were aged 16 years and older; 19.9% vs. 15.4% were 12-15 years). The time-periods with the highest SARS-CoV-2 percent PCR-positivity were Nov 2020-Feb 2021 and July-October 2021 (**Table 1**). From our cohort of 1,782,537 patients, there were 1,260 children with a diagnosis of MIS-C (7.1 cases per 10,000 population), of whom 155 tested positive for SARS-CoV-2.

**Table 1.**
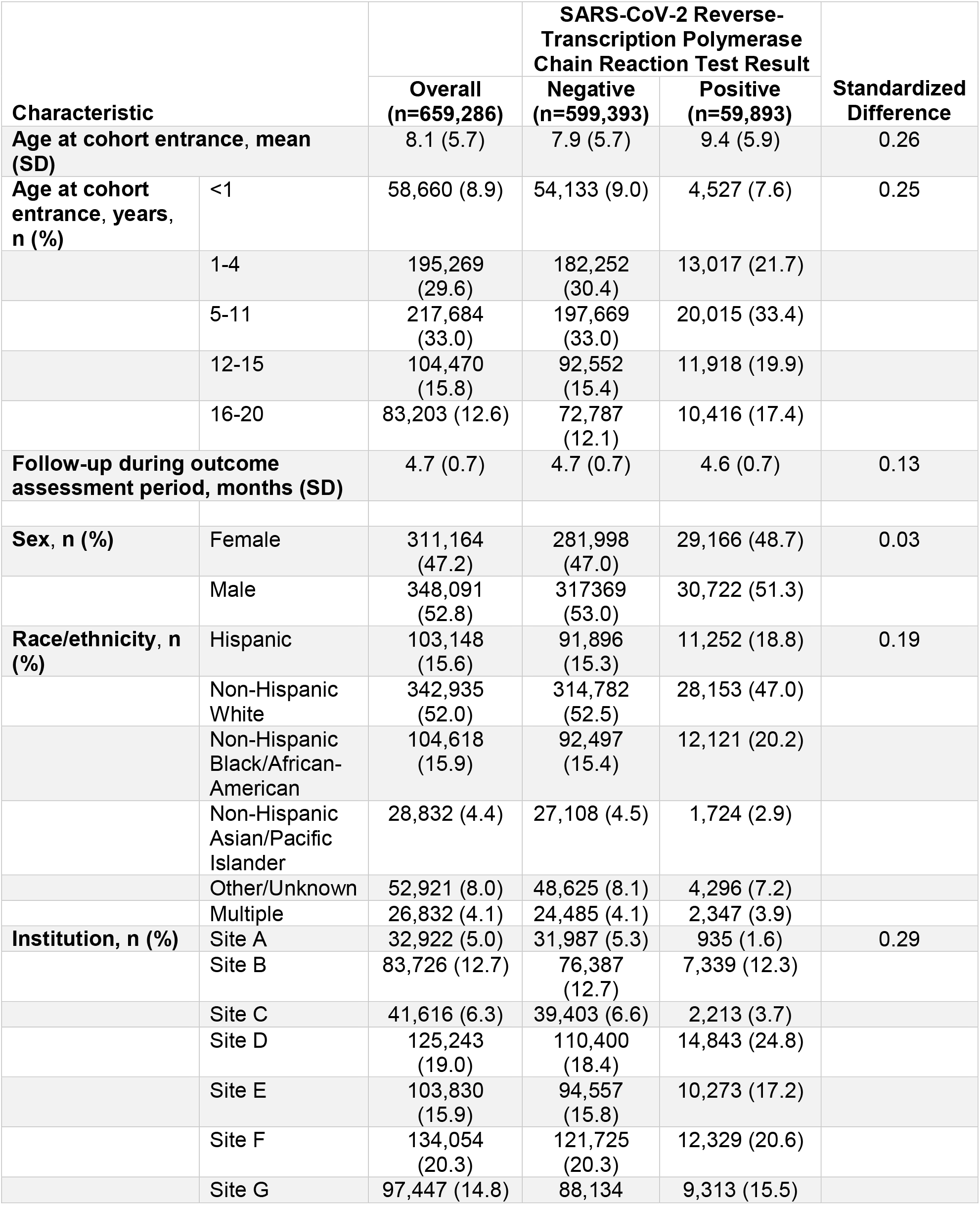
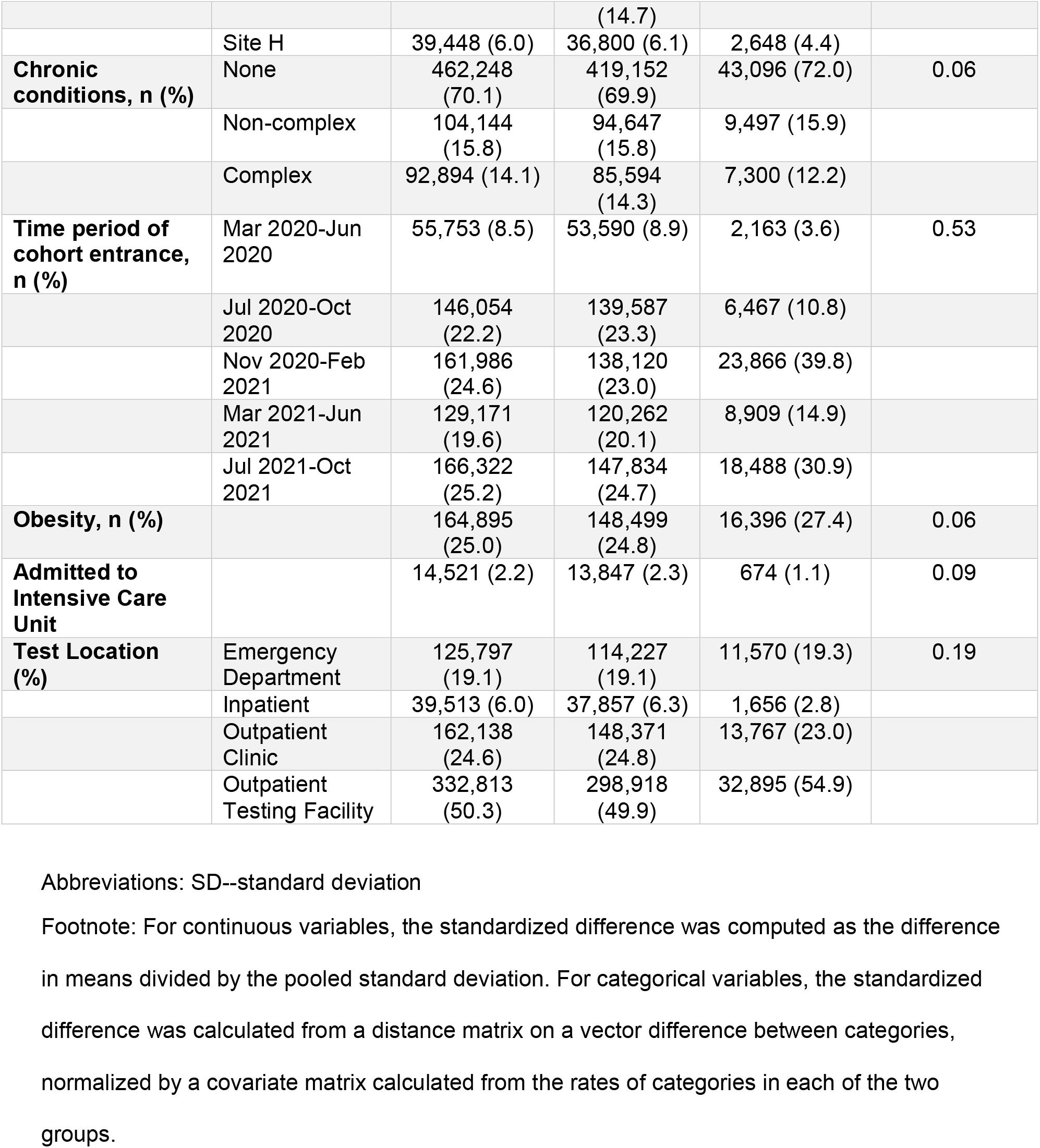
Study sample characteristics.

### Syndromic and Systemic PASC Features

Adjusted analyses of syndromic and systemic features of PASC by cluster category that were clinically-predicted and empirically-supported are summarized in Figures 1 and 2. (Those that were clinically-predicted but not empirically-supported and vice versa are listed in the Supplemental Appendix). During the outcome assessment period (days 28 to 179), PCR-positive children compared with PCR-negative individuals had higher rates of changes in smell and taste (aHR 1.96 [95% CI 1.16-3.32]), loss of smell (aHR 1.85 [95% CI 1.20-2.86]), hair loss (aHR 1.58 [95% CI 1.24-2.01]) chest pain (aHR 1.52 [95% CI 1.38-1.68]), abnormal liver enzymes (aHR 1.50 [95% CI 1.27-1.77]), skin rashes (aHR 1.26 [95% CI 1.15-1.38]), fatigue and malaise (aHR 1.24 [95% CI 1.13-1.35]), fever and chills (aHR 1.22 [95% CI 1.16-1.28]), cardiorespiratory signs and symptoms (aHR 1.20 [95% CI 1.15-1.26]), and diarrhea (aHR 1.18 [95% CI 1.09-1.29]) (Figure 1).

**Figure 1.**
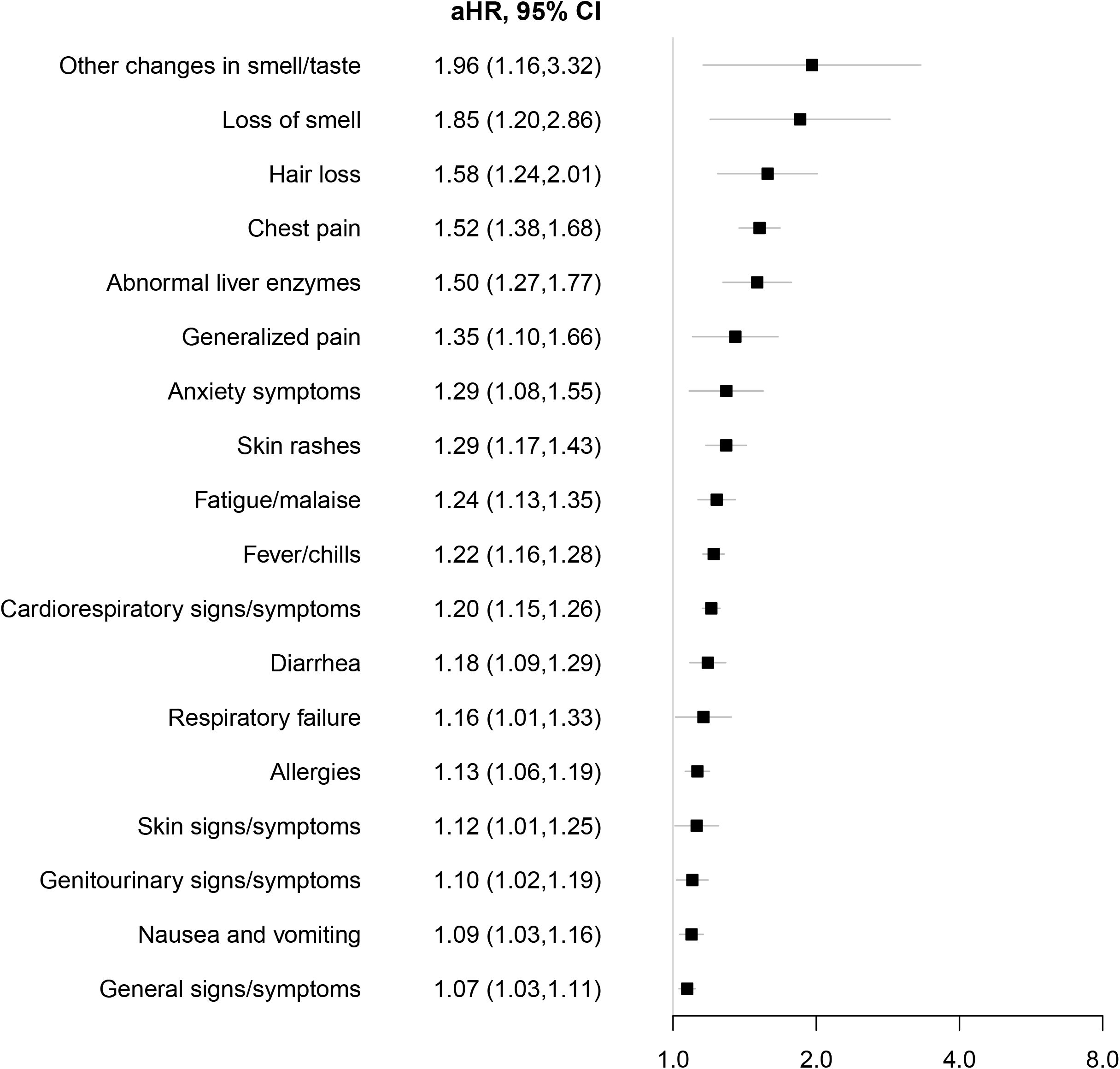
Syndromic PASC features associated with SARS-CoV-2 infection. Adjusted hazard ratios (aHR) with associated 95% CI among patients who tested positive for SARS-CoV-2 infection versus those who tested negative for the risk of each syndromic feature using Cox proportional hazards models. Models were adjusted for age at cohort entrance, sex, race/ethnicity, institution, testing place location, presence of a complex medical condition and date of cohort entrance.

**Figure 2.**
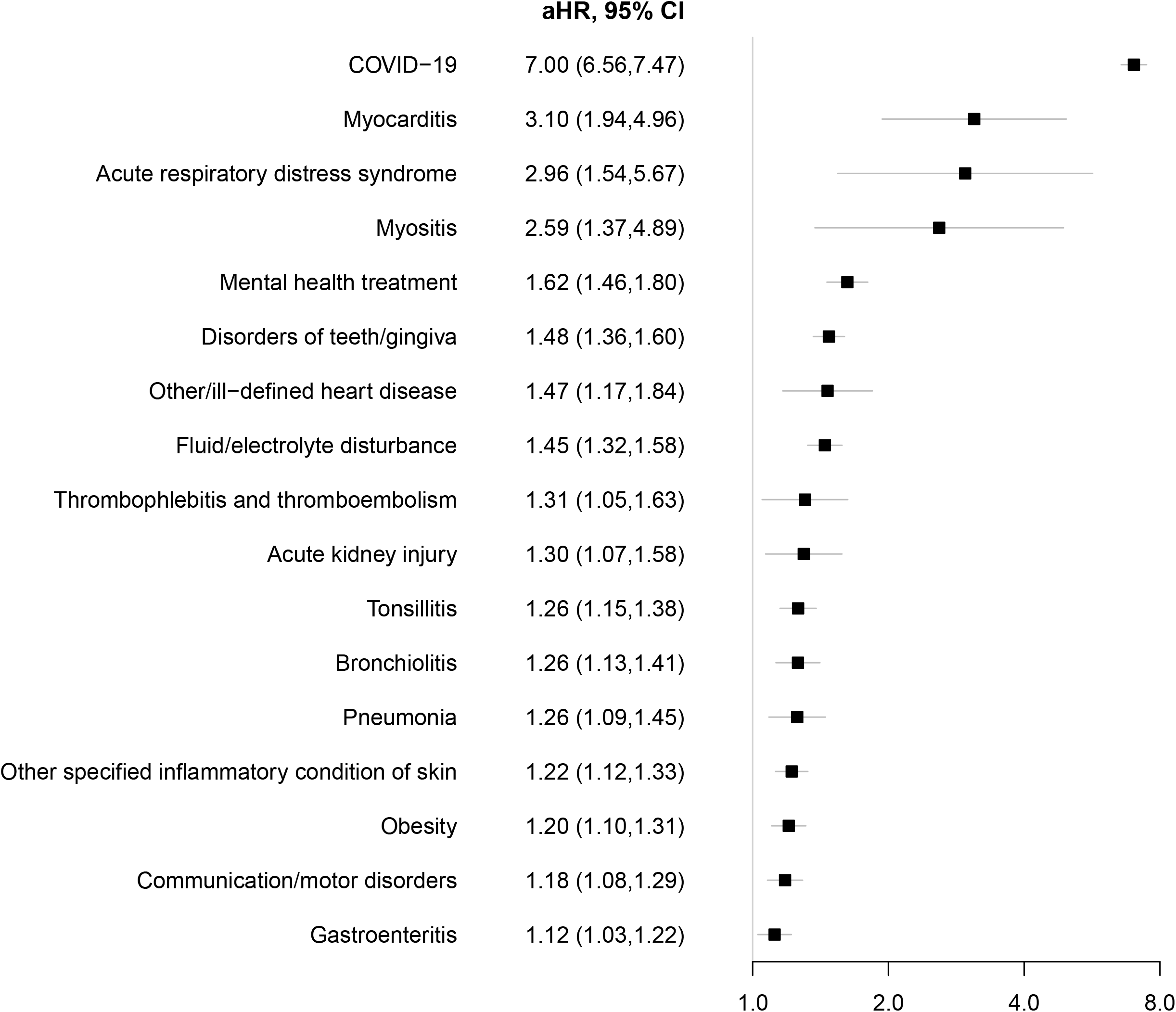
Systemic PASC features associated with SARS-CoV-2 infection. Adjusted hazard ratios (aHR) with associated 95% CI among patients who tested positive for SARS-CoV-2 infection versus those who tested negative for the risk of each systemic feature using Cox proportional hazards models. Models were adjusted for age at cohort entrance, sex, race/ethnicity, institution, testing place location, and date of cohort entrance. For each health condition evaluated, patients with evidence of that condition 18 months before cohort entrance were excluded from the denominator in order to identify incident cases. Each ratio compares the risk of the outcome in children who tested positive for SARS-CoV-2 infection versus those who tested negative. Footnote: The diagnostic cluster for COVID-19 indicates children receiving care for the illness in the post-acute period.

Regarding systemic features, infected cohorts had higher rates of myocarditis (HR 3.10 [95% CI 1.94-4.96]), acute respiratory distress syndrome (aHR 2.96 [95% CI 1.54-5.67]), myositis (aHR 2.59 [95% CI 1.37-4.89]), disorders of teeth/gingiva (aHR 1.48 [95% CI 1.36-1.60]), other/ill-defined heart disease (aHR 1.47 [95% CI 1.17-1.84]), and fluid and electrolyte disturbance (aHR 1.45 [95% CI 1.32-1.58]) (Figure 2). Mental health clusters associated with prior SARS-CoV-2 infection in our analyses were mental health treatment (aHR 1.62 [95% CI 1.46-1.80] and anxiety symptoms (aHR 1.29 [95% CI 1.08-1.55].

### Medications

Medication therapeutic classes clinically-predicted and empirically-supported for their association with COVID-19 illness are shown in the Supplemental Appendix. The five most common were cough and cold preparations, nasal decongestants for systemic use, corticosteroids with antiseptics, opioids, and decongestants.

### Incidence of PASC

The incidence proportions for non-MIS-C related syndromic, systemic and medication features during the 28-179 days after viral testing and their standardized morbidity ratios, adjusted for age and date, of testing are shown in Table 2. The incidence proportion of at least one syndromic, systemic or medication feature of PASC was 41.9% (95% CI 41.4-42.4) in the PCR-positive group and 38.2% (95% CI 38.1-38.4) in those who tested negative for a difference of 3.7% (95% CI 3.2-4.2). The standardized morbidity ratio, which adjusted for differences in the distributions of age and date of cohort entrance between the two groups, was 1.15 (95% CI 1.14-1.17).

**Table 2.** Incidence proportions for syndromic, systemic, and medication features of PASC unrelated to MIS-C observed day 28-179 after viral testing and standardized morbidity ratios adjusted for patient age and date of viral testing.

| PASC Feature | N Incidences |  | N Persons |  | Incidence Proportion (95% CI) |  | Incidence Proportion Difference (95% CI) | Standardized Morbidity Ratio |
| --- | --- | --- | --- | --- | --- | --- | --- | --- |
|  | PCR Test Positive | PCR Test Negative | PCR Test Positive<sup>d</sup> | PCR Test Negative | PCR Test Positive | PCR Test Negative |  |  |
| Syndromic<sup>a</sup> | 10,657 | 108,906 | 54,061 | 548,304 | 19.7% (19.4, 20.0) | 19.9% (19.8, 20.0) | -0.1% (-0.5, 0.2) | 1.07 (1.05, 1.09) |
| Systemic<sup>a</sup> | 7,084 | 50,786 | 52,027 | 529,741 | 13.6% (13.3, 13.9) | 9.6% (9.5, 9.7) | 4.0% (3.7, 4.3) | 1.49 (1.45, 1.52) |
| Medication | 7,871 | 73,651 | 40,742 | 425,741 | 19.3% (18.9, 19.7) | 17.3% (17.2, 17.4) | 2.0% (1.6, 2.4) | 1.19 (1.16, 1.22) |
| Syndromic or systemic<sup>b</sup> | 14,418 | 133,328 | 50,180 | 510,045 | 28.7% (28.3, 29.1) | 26.1% (26.0, 26.3) | 2.6% (2.2, 3.0) | 1.16 (1.14, 1.18) |
| Syndromic, systemic, or medication<sup>c</sup> | 17,498 | 162,228 | 41,756 | 424,215 | 41.9% (41.4, 42.4) | 38.2% (38.1, 38.4) | 3.7% (3.2, 4.2) | 1.15 (1.14, 1.17) |

Abbreviations: PASC- Post-acute sequelae of COVID-19; PCR- polymerase chain reaction; CI- Confidence Interval
<sup>a</sup> Any syndromic and any systemic refer to the features in Figures 1 and 2, respectively, that were clinically predicted or plausible and empirically supported in the proportional hazards model.
<sup>b</sup> Either syndromic or systemic refers to the union of the features from any syndromic or systemic.
<sup>c</sup> Syndromic, systemic or medication refers to the union of the features from any syndromic, systemic or medication.
<sup>d</sup> Number of PCR test positive and negative patients differs across PASC features due to washout of chronic conditions.

### Factors associated with non-MIS-C related PASC

Proportional hazards models for non-MIS-C related PASC, defined by any clinically-predicted and empirically-supported syndromic features, systemic conditions and medication features are shown in Table 3. These analyses demonstrated the highest rate of any PASC feature in children <5 years-old, requiring ICU level care during the acute illness episode, or with a complex chronic condition. The risk of non-MIS-C related PASC was highest during the March-June 2020 time period.

**Table 3.**
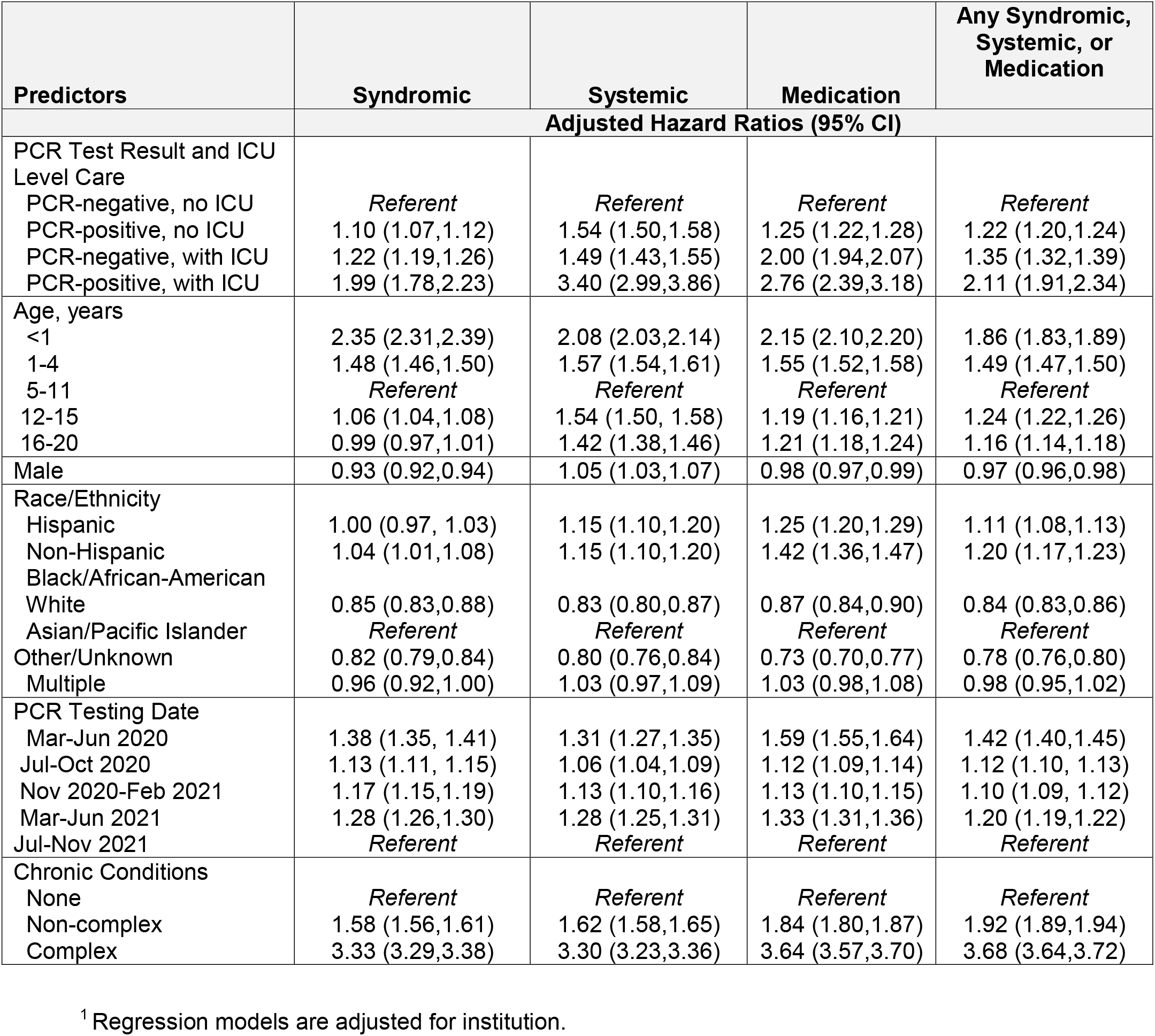
Proportional hazards models for PASC unrelated to MIS-C defined by syndromic features, systemic conditions, medications features, or all three.^1^.

## Discussion

In this exploratory study we identified possible clinical features of PASC including symptoms, conditions and medications to define these features. Our results showed features that have been well-described in adult populations, including changes in taste or smell, chest pain, fatigue/malaise, cardiorespiratory sign/symptoms, and fever/chills. In this pediatric population, however, we also note other features such as abnormal liver enzymes, hair loss, skin rashes, and diarrhea that occurred more commonly in children in the 1 to 6 months after SARS-CoV-2 infection compared with negative patients. Among systemic features, myocarditis was the condition with the strongest association with SARS-CoV-2 infection and is recognized as an important complication in this age group.^30,31^ Myositis, encounters for mental health treatment, and respiratory infections including tonsillitis, pneumonia and bronchiolitis were also associated with COVID-19.

To estimate the burden of PASC unrelated to MIS-C, we computed the incidence proportion difference of clinically-predicted and empirically-supported PASC features between PCR-positive and PCR-negative groups, which was 3.7% (95% CI, 3.2%, 4.2%). This proportion is an estimate of attributable burden of any syndromic, systemic, or medication-related PASC feature in the sample of children we studied. This estimate should be considered preliminary as it is based entirely on diagnosis and medication codes in EHRs, which depend on clinicians’ coding practices and may exclude children who do not seek healthcare for PASC symptoms. Nonetheless, the strength of association of PASC increased with acute COVID-19 severity, similar to previous reports,^32^ and medical complexity.

Early reports of PASC in children reporting higher incidence than in this study were prone to systematic selection biases of cases in the absence of appropriate controls. Patients were recruited from long COVID clinics^17,20^ or online support groups,^33^ with data limited to self-reported symptoms, some without a laboratory-confirmed diagnosis of SARS-CoV-2, limiting the ability to differentiate symptoms and conditions attributable to SARS-CoV-2 infection versus illnesses that may have been exacerbated by the situational context of the pandemic (e.g. delayed access to healthcare, prolonged social isolation). Other studies focused on hospitalized patients,^34^ biasing the sample to patients who were at higher risk of more persistent or severe post-acute symptoms. Our study’s strengths lie in capturing patients with PCR-confirmed infection and including a control group of PCR-negative children, thereby minimizing bias from differences in health-seeking behaviors between test positive and test negative cohorts. Finally, our population includes children from diverse geographical locations across the US, and encompasses ambulatory clinics, outpatient referrals to testing sites, subspecialty clinics as well as hospitalized patients, which provides a wider spectrum of presentations.

Unlike case series that describe the prevalence of PASC features in a cohort of children previously infected, we compared the incidence rate of these symptoms in test positive versus test negative patients to estimate an attributable burden. With this approach, we reported a preliminary estimate of lower incidence of PASC in children compared with adults.^5-9^ Our findings are similar to recent pediatric studies which use population-based approaches, and/or control groups.^15,35,36^ Notably, the frequency of any PASC-related symptoms is high in both PCR-positive and PCR-negative patients (41.9% vs. 38.2%), similar to observations in adult studies.^37,38^ Nonetheless, the attributable and measurable burden of PASC may still differ in children in comparison to adults. Potential reasons for these differences may be under-recognition of signs and symptoms associated with PASC due to a dynamic developmental trajectory, or age-specific differences in the immune response to infection in PASC related either to ongoing viral replication or an aberrant response.

Our findings have limitations that warrant discussion. This EHR-based study identified symptoms, signs, and diagnoses that were significant enough to prompt health service use and be coded by clinicians as a reason for an encounter. Our approach may have missed some findings that may be stored in laboratory, procedural, radiological and other unstructured text data. The true burden of PASC may be underestimated from EHR data, with data from open health systems, with the potential for infrequent follow-up visits to academic centers of excellence for milder symptoms. For this reason, we limited our cohort to at least one visit in the prior three years, to identify active patients within the health system; similar strategies have been employed in other PEDSnet studies of COVID-19;^39,40^ the benefits of such approaches have been reported previously.^41^ However, this may underestimate the burden of PASC by excluding previously healthy children who did not have prior health encounters. Next, our test negative cohort may include individuals with SARS-CoV-2 infection who may have had testing conducted outside our health systems. These limitations may have biased results towards the null. Further, we did not identify specific race/ethnic groups as a risk factor for PASC despite the fact that SARS-CoV-2 disproportionally affects minority communities, which may reflect differences in care seeking behavior and access to care. Finally, children who tested positive may be more likely to seek medical care, however we do not anticipate this to be creating significant bias given our low incident proportion difference, which is similar to findings from other controlled studies.

While there were significant features associated with SARS-CoV-2 infection in our study that overlapped with findings reported in the literature in adults (such as loss of taste and smell^42,43^), several distinctions are worth noting. First, neurological manifestations such as headache, vertigo and paresthesias, which are commonly reported in adults, were not significant findings in our study. In addition, memory loss and “brain fog” have been documented up to 3 months post SARS-CoV-2 infection in adults,^6,7,44^ but were not identified in our study, possibly due to these symptoms not being adequately captured through diagnosis codes. Next, respiratory symptoms such as persistent cough and dyspnea also did not feature as prominently in our study as other observational cohort studies in adults ^9,45-47^ and children,^17,18,20,36^ however, these symptoms were detected through capture of cough and cold preparations in our exploration of medication utilization. Interestingly, during the 1-6 months following infection, our study detected increased rates of pneumonia, tonsillitis and bronchiolitis, which may represent persistent pulmonary symptoms, as well as the potential for an increased susceptibility to other infections from lung pathology developing after SARS-CoV-2 infection. While the mechanisms for injury in patients with pulmonary manifestations of PASC are providing some insights in adults,^48,49^ further research is required to determine how the lung pathophysiology in children differs from adults both in acute COVID-19 infection (for example, increased susceptibility to infection or asthma) and in the mechanisms driving long-term symptoms.

## Conclusion

In the first large-scale study to understand the features of PASC in children, the burden of PASC appeared to be low and systemic features were predominant and varied. Our findings suggest that the burden and risk windows of PASC may differ between children and adults. Future studies, including long term prospective studies, such as NIH RECOVER, are needed to fully elucidate PASC phenotypes.

## Data Availability

All data produced in the present study are available upon reasonable request to the authors

https://github.com/PEDSnet/PASC

## Abbreviations

PASC: post-acute sequelae of SARS-CoV-2 infection
COVID-19: coronavirus disease 2019
SARS-CoV-2: severe acute respiratory syndrome coronavirus 2
PCR: polymerase chain reaction
EHR: electronic health record
aHR: adjusted hazard ratio;
MIS-C: multisystem inflammatory syndrome in children
ED: emergency department
UC: urgent care
CI: confidence intervals
ICD-10: International Classification of Diseases, version 10

## Access to data and data analysis

CF, VL and HR had full access to all the data in the study and take responsibility for the integrity of the data and the accuracy of the data analysis.

