## Supplementary material for "Clinical features and burden of post-acute sequelae of SARS-CoV-2 infection in children and adolescents: an exploratory EHR-based cohort study from the RECOVER program": Online Supplement

**Online-only material**

**eTable 1. Codesets for Systemic and Syndromic Features of PASC and Medication Therapeutic Classes associated with PASC**

**eFigure 1. Medication therapeutic classes associated with PASC.**

**eFigure 2. Kaplan-Meier plot for risk of any syndromic feature of PASC**

**eFigure 3. Kaplan-Meier plot for risk of any systemic feature of PASC**

**eFigure 4. Kaplan-Meier plot for risk of any medication feature of PASC**

**eFigure 5. Kaplan-Meier plot for risk of any syndromic, systemic or medication feature of PASC**

**eTable 1. Codesets for Systemic and Syndromic Features of PASC and Medication Therapeutic Classes associated with PASC^a^**

|  | **Syndromic (symptoms, signs, non-specific laboratory abnormalities)** | **Systemic (diagnosed health conditions)** | **Medication Therapeutic Classes** |
| --- | --- | --- | --- |
| Clinically predicted and empirically supported (i.e., lower limit of the 95% CI for the outcome was greater than 1) | - Other changes in smell/taste - Loss of smell - Hair loss - Chest pain - Abnormal liver enzymes - Generalized pain - Anxiety symptoms - Skin rashes - Fever/chills - Cardiorespiratory signs/symptoms - Fatigue/malaise - Diarrhea - Allergies - Skin signs/symptoms - Nausea and vomiting - General signs and symptoms - Genitourinary signs and symtpoms | - COVID-19 - Multisystem inflammatory syndrome - PASC - Acute respiratory distress syndrome - Myocarditis - Myositis - Mental health treatment - Fluid/electrolyte disturbance - Disorders of teeth/gingiva - Other/ill-defined heart diseasex - Acute kidney injury - Thrombophlebitis and thromboembolism - Pneumonia - Bronchiolitis - Tonsilitis - Other/specified inflammatory condition of skin - Obesity - Communication/motor disorders - Respiratory failure - Gastroenteritis | - Antiarrhythmics, Class I and III - Antiemetics/antinauseants - Antiinflammatory and antirheumatic agents in combination - Cough and cold preparations - Systemic corticosteroids - Corticosteroids with antiseptics - Decongestants - Direct acting antivirals - Hypnotics/sedatives - Opioids - Parasympathomimetics - Propulsives - Cough suppressants and expectorants, combinations - Nasal decongestants for systemic use |
| Clinically predicted but not empirically supported | - Abdominal pain - Abdominal signs and symptoms - Anger and aggression - Anorexia - Anxiety disorder - Arthralgia - Attention symptoms - Balance problems - Bruising and bleeding disorders - Chilblain - Cognitive impairment - Cognitive signs and symptoms - Coma - Constipation - Cough - Delirium - Depressive symptoms - Diseases of the vocal cords and larynx - Dizziness and syncope - Dysphagia - Emotional and behavioral signs and symptoms - Genitourinary signs and symptoms - Headache - Loss of taste - Myalgia - Nasal congestion - Neuraliga and neuritis - Ocular pain - Otalgia - Pruritis - Psychological symptoms, other - Sleep-wake disorders - Speech signs and symptoms - Stress - Visual disturbances - Weight loss | - Academic developmental disorder - Addison disease - Alopecia areata - Antiphospholipid syndrome - Aplastic anemia - Appendicitis - Arrhythmias - Arthritis - Asthma - Attention deficit hyperactivity disorder - Autism spectrum disorder - Autoimmune thyroiditis - Avoidant and restrictive food intake - Bipolar disorder - Bronchitis - Cardiomyopathy - Catatonia - Celiac disease - Chronic kidney disease - Chronic obstructive pulmonary disease and bronchiectasis - Conduct disorder - Dermatitis herpetiformis - Developmental delay - Diabetes mellitus - Diabetes mellitus, type 1 - Diabetes mellitus, type 2 - Dysautonomia - Encopresis - Feeding and eating disorders - Heart disease, other - Hypertension - Hypothyroidism - Idiopathic thrombocytopenic purpura - Inflammatory polyneuropathy - Intellectual disability - Kawasaki disease - Major depression - Minor depression - Neonatal digestive and feeding disorders - Oppositional defiant disorder - Pericarditis - Psoriasis - Psychotic disorder - Pulmonary fibrosis - Rhabdomyolysis - Schizoaffective disorder - Schizophrenia - Scleroderma - Seizures and epilepsy - Sexual dysfunction - Sjogren syndrome - Systemic lupus and other connective tissue disorders - Takotsubo cardiomyopathy - Temporal arteritis - Thyroiditis - Tic disorder - Vitiligo | - ACE Inhibitors, plain - Adrenergics, inhalants - Antidepressants - Antiepileptics - Antihistamines for systemic use - Antiinflammatory agents - Antimigraine preparations - Antithrombotic agents - Beta blocking agents - Drugs for peptic ulcer and gastro-oesophageal reflux disease (GORD) - Immunoglobulins - Immunosuppressants - Other drugs for obstructive airway diseases, inhalants - Expectorants, excl. combinations with cough suppressants - Other mineral supplements |
| Empirically supported but not clinically predicted or evaluated as clinically plausible | - Superficial injury; contusion | - Breast cancer - Neurogenic/neuropathic arthropathy - Personal/family history of disease - Septicemia - Mental health treatment - Adverse effects of drugs and medicaments - Newborn status - Disorders of teeth and gingiva - Influenza - Other and ill-defined heart disease - Complication of device, implant, or graft - Contact dermatitis - Intestinal infection - Parasitic disease - Other injuries - Bacterial infections - Diseases of white blood cells - Tonsillitis - Coagulation and hemorrhagic disorders - Intestinal obstruction and ileus - Postprocedural or postoperative complications - Other specified encounters and counseling - Hypotension - Other specified inflammatory condition of skin - Viral infection - Obesity - Pleurisy, pleural effusion and pulmonary collapse - Other specified status - Complication of other surgical or medical care injury - Other specified upper respiratory infections - Inflammatory conditions of male genital organs - Urinary tract infections - Other specified and unspecified skin disorders - Skin and subcutaneous tissue infections - Administrative encounter - Nervous system pain and pain syndromes - Otitis media | - Ultrasound contrast media - Allergens - Irrigating solutions - Anesthetics, local - Ectoparasiticides, incl. scabicides - Anesthetics, general - Antiinflammatory/antirheumatic agents in combination - Antispasmodics in combination with analgesics - Corticosteroids, combinations with antibiotics - Antiinfectives and antiseptics, excl. combinations with corticosteroids - Bacterial vaccines - Vitamin k and other hemostatics - Other antibacterials - Antiinflammatory agents and antiinfectives in combination - Throat preparations - Bacterial and viral vaccines, combined - Corticosteroids, other combinations - Anti-acne preparations for topical use - Antifibrinolytics - Other otologicals - Antibiotics for topical use - Ascorbic acid (vitamin C), incl. combinations - Blood and related products - Medicated dressings - Local anesthetics - All other therapeutic products - Vitamin B12 and folic acid - Viral vaccines - Antispasmodics in combination with psycholeptics - Muscle relaxants, peripherally acting agents - Antifungals for topical use - Antiseptics and disinfectants - Other ophthalmologicals - Corticosteroids and antiinfectives in combination - Drugs for functional gastrointestinal disorders - I.V. solution additives - Other beta-lactam antibacterials - Beta-lactam antibacterials, penicillins |
| Clinically predicted but outcome found to be of greater risk in test negative cohort (upper limit of 95% CI for outcome was less than 1) | - Balance problems - Stress - Cough - Chilblain - Abdominal pain | - Feeding and eating disorders - Seizures and epilepsy - Major depression - Minor depression - Celiac disease - Dysautonomia - Autism spectrum disorder - Intellectual disability | - ACE inhibitors, plain - Antidepressants - Antiepileptics - Antithrombotic agents - Beta blocking agents - Drugs for peptic ulcer and gastro-oesophageal reflux disease (GORD) - Immunosuppressants |

^a^ Codesets were developed from ICD codes, the full list of the code assignments to clusters is publicly available at [https://github.com/PEDSnet/PASC](https://nam10.safelinks.protection.outlook.com/?url=https%3A%2F%2Fgithub.com%2FPEDSnet%2FPASC&data=04%7C01%7CFORRESTC%40chop.edu%7Cc9d245c53f024ec9adda08d9e6acb412%7Ca611241607b041a59bb1d146b575c975%7C1%7C0%7C637794451691254192%7CUnknown%7CTWFpbGZsb3d8eyJWIjoiMC4wLjAwMDAiLCJQIjoiV2luMzIiLCJBTiI6Ik1haWwiLCJXVCI6Mn0%3D%7C3000&sdata=hQ1grZnE%2FcybQ%2BHlLNfKlSUpDLbHSd7fXLqYYkLhIIo%3D&reserved=0).

**eFigure 1. Medication therapeutic classes associated with PASC**

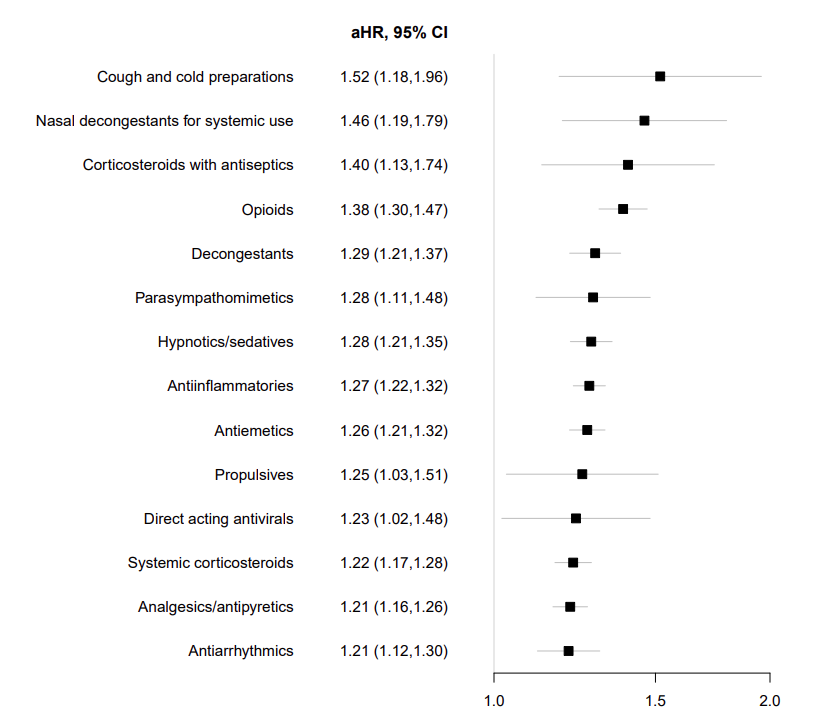

**eFigure 2. Kaplan-Meier plot for any syndromic feature of PASC**

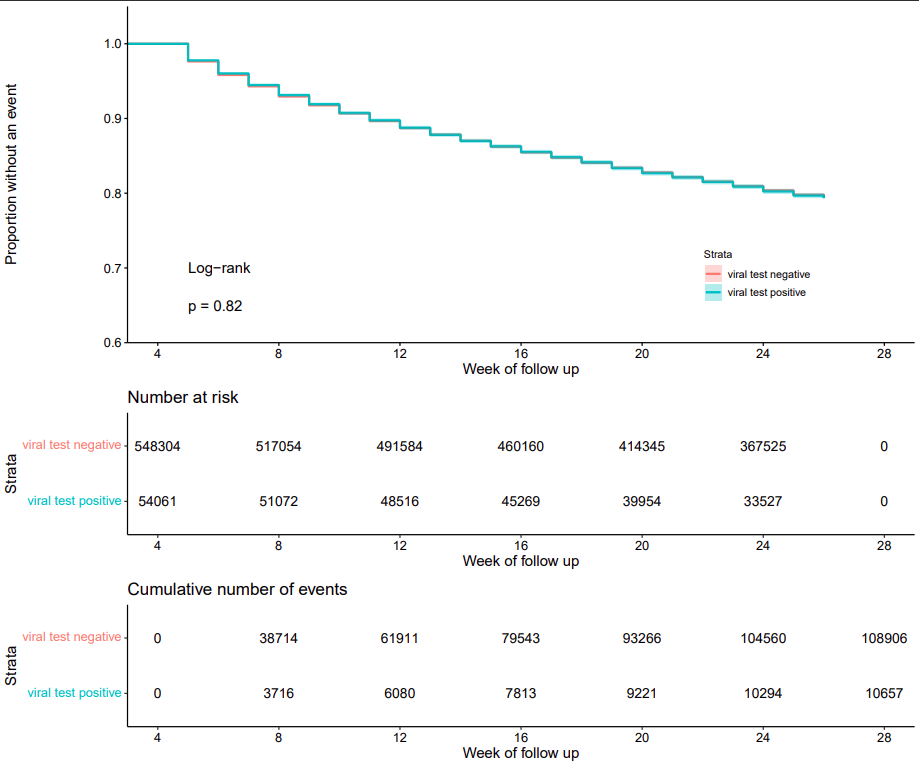

**eFigure 3. Kaplan-Meier plot for any systemic feature of PASC**

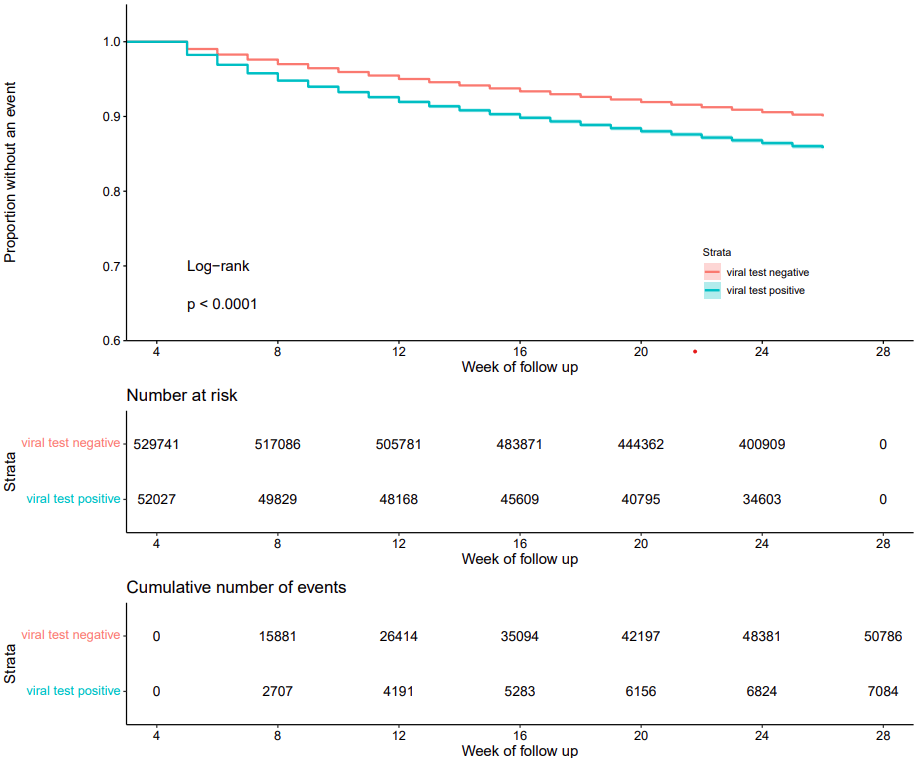

**eFigure 4. Kaplan-Meier plot for any medication feature of PASC**

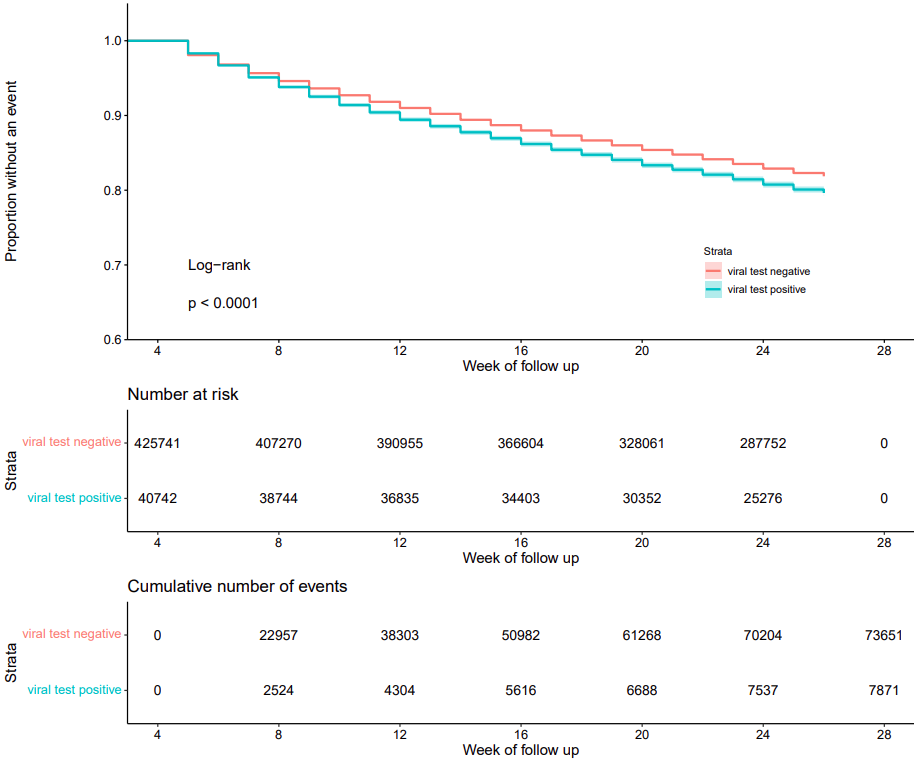

**eFigure 5. Kaplan-Meier plot for any syndromic, systemic or medication feature of PASC**

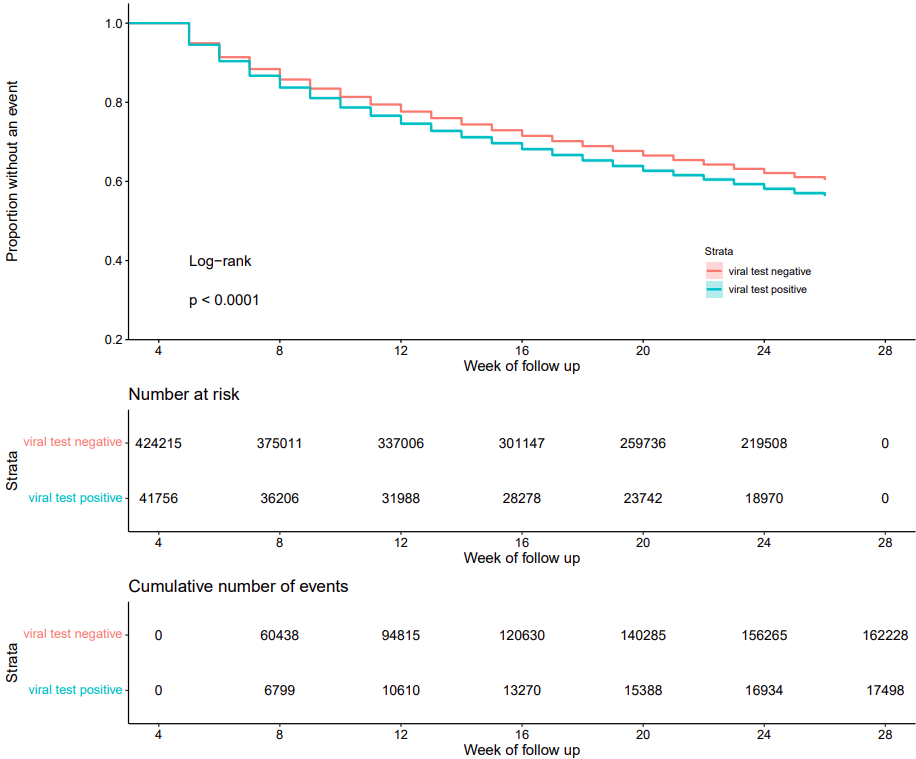
